# Multimodal smoking cessation treatment combining transcranial magnetic stimulation, cognitive behavioral therapy, and nicotine replacement therapy in veterans with posttraumatic stress disorder: A feasibility randomized controlled trial protocol

**DOI:** 10.1101/2023.09.06.23294958

**Authors:** Jonathan R. Young, Jeffrey T. Galla, Carri S. Polick, Zhi-De Deng, Moritz Dannhauer, Angela Kirby, Michelle Dennis, Claire W. Papanikolas, Mariah K. Evans, Scott D. Moore, Eric A. Dedert, Merideth A. Addicott, Lawrence G. Appelbaum, Jean C. Beckham

**Author notes:** Corresponding Author Jonathan R. Young, MD Department of Psychiatry & Behavioral Sciences Duke University School of Medicine, 200 Trent Drive, Box 3620, Durham, NC 27710.

## Abstract

Tobacco-related deaths exceed those resulting from homicides, suicides, motor vehicle accidence, alcohol consumption, illicit substance use, and acquired immunodeficiency syndrome (AIDS), combined. Amongst U.S. veterans, this trend is particularly concerning given that those suffering from posttraumatic stress disorder (PTSD)—about 11% of those receiving care from the Department of Veterans Affairs (VA)—have triple the risk of developing tobacco use disorder (TUD). The most efficacious strategies being used at the VA for smoking cessation only result in a 23% abstinence rate, and veterans with PTSD only achieve a 4.5% abstinence rate. Therefore, there is a critical need to develop more effective treatments for smoking cessation. Recent studies have revealed the insula as integrally involved in the neurocircuitry of TUD, specifically showing that individuals with brain lesions involving this region had drastically improved quit rates. Some of these studies show a probability of quitting up to 5 times greater compared to non-insula lesioned regions). Altered activity of the insula may be involved in the disruption of the salience network’s (SN) connectivity to the executive control network (ECN), which compromises that patient’s ability to switch between interoceptive states focused on cravings to executive and cognitive control. Thus, we propose a feasibility phase II randomized controlled trial (RCT) to study a patterned form of repetitive transcranial magnetic stimulation (rTMS), intermittent theta burst stimulation (iTBS), at 90% of the subject’s resting motor threshold (rMT) applied over a region in the right post-central gyrus most functionally connected to the right posterior insula. We hypothesize that by increasing functional connectivity between the SN with the ECN to enhance executive control and by decreasing connectivity with the default mode network (DMN) to reduce interoceptive focus on withdrawal symptoms, we will improve smoking cessation outcomes. Fifty eligible veterans with comorbid TUD and PTSD will be randomly assigned to two conditions: active-iTBS + cognitive behavioral therapy (CBT) + nicotine replacement therapy (NRT) (*n*=25) or sham-iTBS + CBT + NRT (*n*=25). The primary outcome, feasibility, will be determined by achieving a recruitment of 50 participants and retention rate of 80%. The success of iTBS will be evaluated through self-reported nicotine use, cravings, withdrawal symptoms, and abstinence following quit date (confirmed by bioverification) along with evaluation for target engagement through neuroimaging changes, specifically connectivity differences between the insula and other regions of interest.

## Introduction

In the United States (US), an estimated 28.3 million adults currently smoke cigarettes [1], which causes death or disability in approximately half of this population [2]. There are more tobacco-related deaths than those related to other causes of mortality including acquired immunodeficiency syndrome (AIDS), illicit substance use, alcohol consumption, motor vehicle accidents, suicides and homicides combined [3]. Additionally, individuals with mental illness are disproportionately affected by the medical burden of tobacco use [4]. Smoking rates are especially high among US veterans, and those with posttraumatic stress disorder (PTSD) are more likely to continue smoking despite treatment, increasing the risk of poor health outcomes [5] and reduced life expectancy [6]. Despite substantial progress in developing smoking cessation interventions for veterans with PTSD [7, 8], many veterans continue to smoke and are in critical need of innovative treatment options.

The most efficacious strategies found in VA smoking cessation clinics combine pharmacotherapy with multiple formats of cognitive-behavioral interventions, including self-help materials, group therapy, telephone counseling, and pharmacotherapy, resulting in an abstinence rate of just 23% [9]. Clinic attendance rates at specialty smoking cessation clinics are approximately 13%–14% for all veterans, and those with PTSD are even less likely to follow through on treatment referral [10]. Further, among veterans with PTSD who *do* present to smoking cessation clinics, the efficacy of interventions is even lower [10]. This was demonstrated in the largest smoking cessation trial in veterans with PTSD, with a biochemically verified [bio-verified; exhaled carbon monoxide (CO) < 8 parts per million (ppm) or urine cotinine < 100 nanograms per milliliter (ng/mL)] cessation rate of only 4.5% at 12 months [10]. These findings underscore the urgent need to innovate and develop more efficacious smoking cessation interventions for this clinical population.

Recently, a repetitive transcranial magnetic stimulation (rTMS) coil (H4-coil, BrainsWay, Israel) designed to broadly stimulate bilateral prefrontal cortices and insula received US regulatory clearance as a short-term smoking cessation aid following a positive multicenter clinical trial [11–13]. Each rTMS session was preceded by a brief provocation procedure to induce nicotine craving. However, most clinical trials (including this pivotal study) on rTMS for smoking cessation have been conducted in civilian patients and have excluded individuals with psychiatric conditions. To improve smoking cessation treatment options for veterans with PTSD, it is critical to evaluate novel brain stimulation methods such as rTMS in this vulnerable population. Furthermore, the development of neuroscience-informed techniques to enhance rTMS, such as neuronavigation based on resting-state functional magnetic resonance imaging (rs-fMRI), is important, allowing clinicians and researchers to not only provide individualized rTMS for smoking cessation but also better understand its mechanisms of action.

This study will use state-of-the-art TMS techniques: 1) intermittent theta-burst stimulation (iTBS), 2) accelerated treatment schedule, 3) individualized fMRI-guided targeting with neuronavigation, and 4) synergism with behavioral pharmacologic interventions. First, the approved rTMS protocol uses 10 Hz rTMS. Depression studies indicate that iTBS is as efficacious as high-frequency rTMS while having the advantage of faster treatment time, offering greater efficiency and potentially greater patient tolerability [14]. Second, the rTMS for smoking cessation protocol approved by the U.S. Food and Drug Administration (FDA) involves once-daily treatment sessions. Accelerated TMS involves administering TMS for several sessions per day, which may help produce a more rapid and robust response in a shorter amount of time [15]. Third, the development of neuroscience-informed techniques to enhance TMS, such as neuronavigation based on resting-state functional magnetic resonance imaging (rs-fMRI), is critical for individualizing TMS for smoking cessation and understanding mechanisms of action. Our group has previously demonstrated the ability to non-invasively modulate the activity of deep brain structures, including the insula, by targeting a functionally connected cortical target with rTMS [16]. Finally, combining TMS with evidence-based CBT and NRT has the potential to further enhance the effectiveness of smoking cessation treatment. To date, collected data suggest that the proposed intervention—neuronavigated rTMS over an individualized target within the right post-central gyrus, informed by rs-fMRI, in combination with CBT and NRT—is both feasible and acceptable for veterans with PTSD seeking smoking cessation [17].

The long-term goal of this study is to optimize smoking cessation for veterans with PTSD by further developing a multimodal smoking cessation intervention that adds innovative TMS to evidence-based CBT and NRT. To our knowledge, this will be the first study to evaluate the efficacy of neuronavigated rTMS over a region functionally connected to the insula through a multimodal intervention approach combining the evidence-based treatments of cognitive behavioral counseling for smoking cessation and NRT. The timing of this trial is optimal given the national expansion of rTMS clinical services at VA hospitals via the National Clinical rTMS Program. Due to this infrastructure, the proposed treatment approach could ultimately be adopted at VA hospitals throughout the country. Aligning with the shift to improve reproducibility in neuroimaging research, the aim of this protocol paper is to increase transparency regarding study methodology [18].

## Methods

### Study Design

The proposed study is a two-arm, parallel design, double-blinded, sham-controlled, randomized trial comparing active-vs sham-iTBS neuronavigated to a region on the post-central gyrus functionally connected to the insula in addition to evidence-based CBT for smoking cessation and NRT (see **Figure 2**). Fifty eligible participants will be randomized to one of two conditions: active-iTBS + CBT + NRT (*n*=25) or sham-iTBS + CBT + NRT (*n*=25). Participants will receive baseline structural and functional magnetic resonance imaging (fMRI), twice-daily neuronavigated active or sham iTBS treatments for five consecutive days in the week prior to their intended quit date, followed by a post-treatment MRI. Each iTBS session consists of 1800 pulses delivered at 90% resting motor threshold (rMT) at cortex; the two daily sessions are separated by 50 minutes. All participants will receive 5 weekly sessions of CBT for smoking cessation, starting in the week prior to iTBS. Session 2 occurs during the week of iTBS, and session 3 in the week immediately following iTBS. CBT session 3 is designated as the participant’s intended quit date. All participants will also receive prescriptions of nicotine replacement in the form of 21 mg, 14 mg, and/or 7 mg patches and 2–4 mg nicotine gum or lozenges starting on the quit date and continuing for up to 90 days at individually tapered doses.

**Figure 1.**
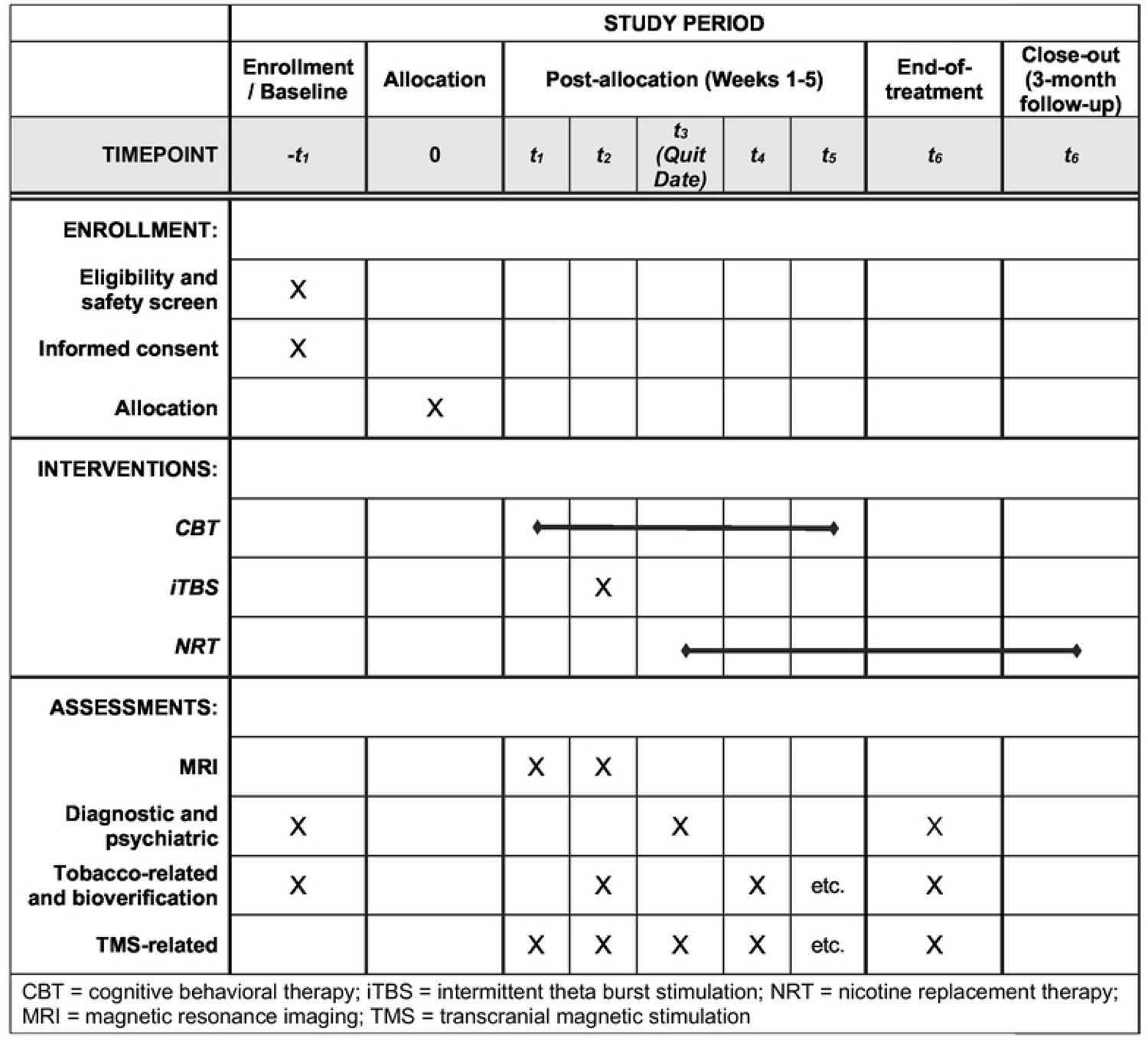
Schedule of study enrollment, interventions, and assessments.

**Figure 2.**
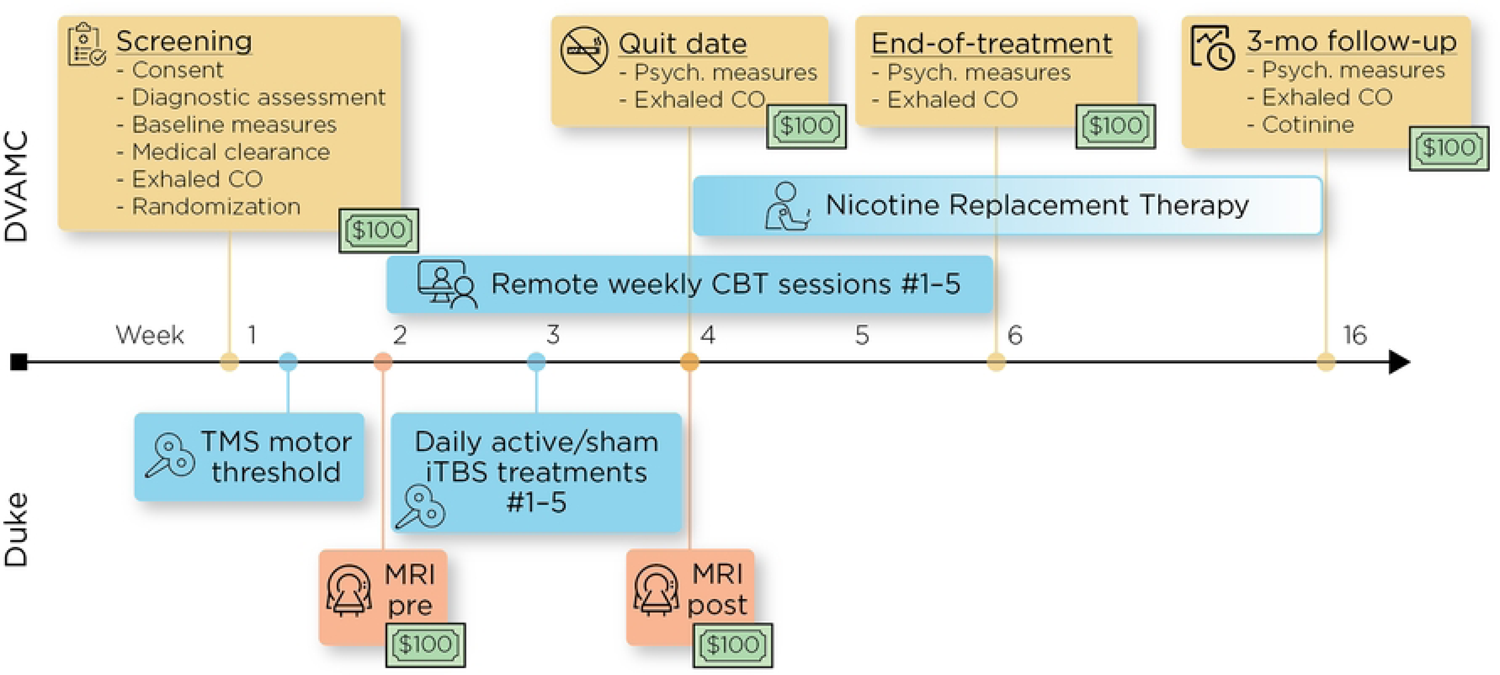

The results of this study will address one primary and two secondary aims: (1) to evaluate feasibility and acceptability of the treatment protocol, (2) to estimate effect size of the iTBS intervention, and (3) to demonstrate neural target engagement using fMRI. Feasibility will be determined through recruitment and retention of participants, with a target enrollment of 50 participants and an 80% retention rate (40 completers). Acceptability will be determined by average scores of at least 70% on a 10-point acceptability measure. Efficacy will be evaluated by estimating the effect size via a 7-day point prevalence abstinence at the end-of-treatment and 3-months post-quit date using both self-reported cigarette use and bioverification. Target engagement on fMRI will be determined by evaluating between-group differences on functional connectivity changes between the stimulation target at the sensorimotor cortex and right posterior insula. Additional analyses will evaluate network-level changes on resting-state functional connectivity (rsFC) using the right posterior insula as a region-of-interest; changes in nicotine dependence using the Fagerström Test for Nicotine Dependence (FTND) [19]; changes in nicotine cravings using the Brief Questionnaire of Smoking Urges [20]; changes in nicotine withdrawal on the Minnesota Nicotine Withdrawal Scale-Revised (MNWS-R) [21]; changes in PTSD symptoms as measured by the PTSD Checklist for DSM-5 (PCL-5) [22], changes in depressive symptoms as measured by the Inventory of Depressive Symptoms-Self Report (IDS-SR) [23]; changes in alcohol use as measured by the alcohol use disorders identification test-consumption (Alcohol Use Disorders Identification Test (AUDIT-C) [24]) screening tool; and changes in illicit substance use as measured by the Drug Abuse Screening Test ([25]).

### Participants and Recruitment

Participants will be recruited at the Durham VA Health Care System through proactive methods (e.g., mailings, flyers) and clinical referrals. Durham, NC, and surrounding areas are heavily populated by veterans. Over 9,000 Afghanistan/Iraq veterans are enrolled at the Durham VA Health Care System (DVAHCS), and clinical reminder data indicate that 1 in 3 (32.7%) are currently smoking. Participants will receive reimbursement for their time regardless of study completion, consistent with the extent of participation, up to $600 ($100 per non-treatment session). Study visit payments are to compensate participants for their time and travel expenses. Based on prior feasibility trials [16, 17] we estimate that approximately 120 veterans will be consented and screened, 50 will be enrolled and randomized to a study condition, and a final sample of 40 participants will complete at least 90% of study procedures. As the overall goal of the proposed study is to test feasibility and begin to estimate effect sizes of the intervention and study procedures, the proposed sample size for enrollment (*n*=50) should provide adequate data for analysis and interpretation (*n*=40).

We will use VA Data Access Request Tracker (DART) requests to identify additional veterans with PTSD and tobacco use disorders. We have used similar procedures to successfully recruit veterans with PTSD in the past. Any potential participant who is identified via a DART request will be sent a recruitment letter by mail or e-mail that provides basic information about the study. The letter will inform veterans that they will be contacted by phone in the coming days regarding their interest in participating in the study. In the letter, potential participants will be given an “opt-out” number to call to decline participation and/or further contact regarding participation. One to two weeks after the mailing, veterans who have not called to decline participation will be called by a study staff member to request their participation in the research study.

If any participant contacts or is contacted by the study coordinator regarding participation, a telephone script will be used to inform them about the study and conduct a preliminary determination of eligibility. If after the telephone screening a participant is considered potentially eligible for participation, he/she will be scheduled for an initial screening session. Once a participant reports to the laboratory to begin the study, the study staff member obtaining consent will explain the study in detail, provide the participant with a DVAHCS IRB-approved written consent form explaining the procedures and risks, and answer any questions. The initial consent process and documentation takes place either in a quiet, private office at DVAHCS or remotely using DocuSign. Participants will be encouraged to read the consent prior to participation. Participants will also be given a copy of the signed informed consent form and phone numbers to call if they have additional questions about the consent form or the research, if they have any problems during the study, or if they have questions about participating in research studies in general. No study procedures will begin until informed consent has been obtained. Participants deemed to be eligible at DVAHCS will subsequently complete a separate consent form for procedures at Duke University Medical Center (DUMC) (see **Appendix 1** for a sample DVAHCS consent form and **Appendix 2** for a sample DUMC consent form).

Eligible participants will be US veterans, ages 18–75, who meet DSM-5 criteria for tobacco use disorder (TUD) and PTSD diagnoses, have been smoking ≥10 cigarettes daily on average over the past 6 months with baseline CO > 6 ppm, are willing to make a smoking cessation attempt, speak and write fluent conversational English, and are appropriate candidates for study procedures (i.e., absence of contraindications for MRI, TMS, and NRT) (**Table 1**).

**Table 1.** Study eligibility criteria.

| Inclusion | Exclusion |
| --- | --- |
| • US veteran<br>• 18-75 years old<br>• Meet DSM-5 criteria for current TUD & PTSD<br>• Smoking >10 cigarettes/day on average for past 6 months, with carbon monoxide (CO) level > 6 ppm<br>• Willing to make cessation attempt<br>• Speaks, reads and writes English<br>• Willing and able to provide informed consent<br>• Has been stable on psychotropic medications for at least three months | • Has had a substance use disorder other than tobacco in the preceding 3 months<br>• Has a history of myocardial infarction in the past 6 months or has another contraindication to NRT<br>• Has a contraindication to TMS or MRI<br>○ Personal or family history of a seizures or epilepsy<br>○ History of neurological condition that increases the risk of seizures including stroke or transient ischemic attack, cerebral aneurysm, or severe traumatic brain injury from a penetrating head injury, loss of consciousness > 20 minutes at time of traumatic injury, requiring an anticonvulsant medication for seizures, and/or found to have encephalomalacia on baseline MRI<br>○ Structural brain lesion, or prior brain surgery<br>○ Ferromagnetic metal in head (including shrapnel)<br>○ Implanted devices that may be affected by MRI or TMS (pacemaker, medication pump, cochlear implant, implanted deep brain stimulator)<br>○ Is pregnant<br>• Is unable to complete study procedures<br>• Is currently prescribed bupropion and/or varenicline<br>• Uses other forms of nicotine such as cigars, pipes, chewing tobacco, or vaping<br>• Is unable to provide informed consent due to a major neurocognitive disorder or other reason<br>• Meets criteria for a primary psychotic disorder or current manic episode<br>• Is currently imprisoned or psychiatrically hospitalized<br>• Has previously received rTMS |

Participants may be on stable psychotropic medications, including standard medications for PTSD, but the study team will discourage changes during the trial. To avoid potential confounding effects on smoking cessation as well as potential for modifying seizure risk during rTMS [26, 27], veterans who are taking either bupropion or varenicline will be excluded.

Participants will be excluded from the study if they have contraindications to TMS. These include any conditions that increase the risk of seizures, including increased intracranial pressure, neurological conditions (e.g., epilepsy or history of seizures, stroke, space-occupying brain lesions), implanted device or any metal in head or neck except related to dental work; or if they expect to be unstable on their medication regimen during the study. Additionally, we will exclude participants who have a history of myocardial infarction in the past 6 months or other contraindication to NRT; use other forms of nicotine such as cigars, pipes, chewing tobacco or vaping; are pregnant or not willing to use contraception (for women of child bearing potential); are currently enrolled in another smoking cessation trial; have major neurocognitive disorder or other reason to not have capacity for informed consent; meet the Structured Clinical Interview for DSM-5 (SCID-5) criteria for a primary psychotic disorder, current mania, or substance use disorder other than nicotine/tobacco in the preceding 3 months; have moderate to severe traumatic brain injury (including history of penetrating head injury, loss of consciousness >20 minutes at time of traumatic injury); require anticonvulsant medications; show encephalomalacia on baseline MRI; or are currently imprisoned or psychiatrically hospitalized. Participants may voluntarily withdraw from participation at any time. The study staff may withdraw a participant one or more of the following reasons: failure to follow the instructions of the study staff, inability to complete the study requirements, or inability to reach participant by telephone after multiple attempts.

### Assessment

Due to COVID-19, and the growing trend of tele-health to increase equity and access, psychiatric assessments will be completed remotely *via* secure platforms. Participants will be initially screened for possible eligibility by phone and then receive a psychiatric interview [SCID-5 and Clinician-Administered PTSD Scale for DSM-5 (CAPS-5)]. Safety screening will be completed in person using the TMS Adult Safety Screen (TASS) [28], the Duke MRI Safety Screening form, and the Ohio State University-Traumatic Brain Injury (OSU-TBI) Identification Method-Short Form. Participants will also complete self-report measures to evaluate for nicotine dependence and cravings, depressive symptoms, and PTSD symptoms. Study inclusion and exclusion criteria will be used to determine eligibility. **Table 2** outlines study measures at major time points.

**Table 2.**
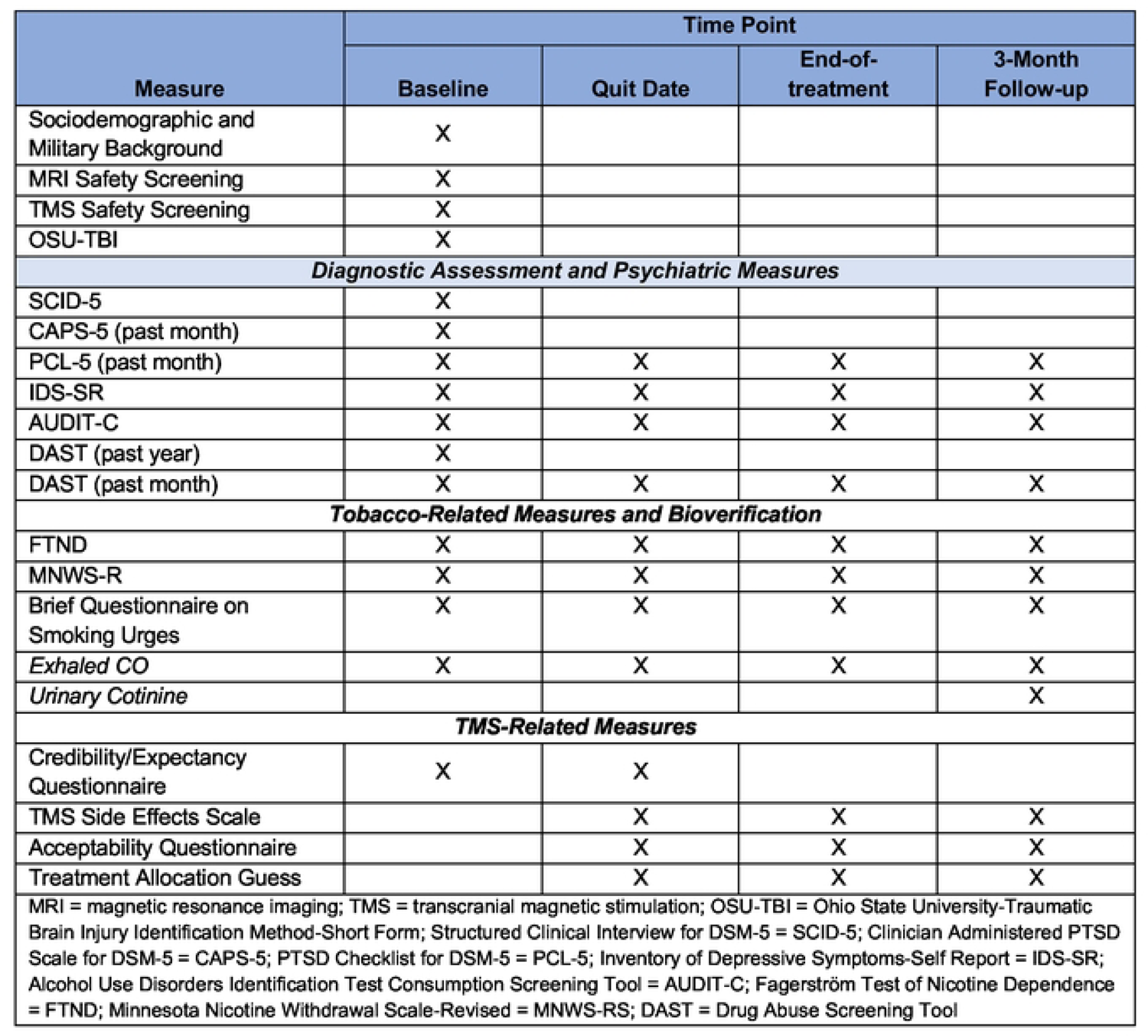
Measures Administered at Major Study Time Points.

| Measure | Time Point |  |  |  |
| --- | --- | --- | --- | --- |
|  | Baseline | Quit Date | End-of-treatment | 3-Month Follow-up |
| Sociodemographic and Military Background | X |  |  |  |
| MRI Safety Screening | X |  |  |  |
| TMS Safety Screening | X |  |  |  |
| OSU-TBI | X |  |  |  |
| **Diagnostic Assessment and Psychiatric Measures** |  |  |  |  |
| SCID-5 | X |  |  |  |
| CAPS-5 (past month) | X |  |  |  |
| PCL-5 (past month) | X | X | X | X |
| IDS-SR | X | X | X | X |
| AUDIT-C | X | X | X | X |
| DAST (past year) | X |  |  |  |
| DAST (past month) | X | X | X | X |
| **Tobacco-Related Measures and Bioverification** |  |  |  |  |
| FTND | X | X | X | X |
| MNWS-R | X | X | X | X |
| Brief Questionnaire on Smoking Urges | X | X | X | X |
| Exhaled CO | X | X | X | X |
| Urinary Cotinine |  |  |  | X |
| **TMS-Related Measures** |  |  |  |  |
| Credibility/Expectancy Questionnaire | X | X |  |  |
| TMS Side Effects Scale |  | X | X | X |
| Acceptability Questionnaire |  | X | X | X |
| Treatment Allocation Guess |  | X | X | X |
| MRI = magnetic resonance imaging; TMS = transcranial magnetic stimulation; OSU-TBI = Ohio State University-Traumatic Brain Injury Identification Method-Short Form; Structured Clinical Interview for DSM-5 = SCID-5; Clinician Administered PTSD Scale for DSM-5 = CAPS-5; PTSD Checklist for DSM-5 = PCL-5; Inventory of Depressive Symptoms-Self Report = IDS-SR; Alcohol Use Disorders Identification Test Consumption Screening Tool = AUDIT-C; Fagerström Test of Nicotine Dependence = FTND; Minnesota Nicotine Withdrawal Scale-Revised = MNWS-RS; DAST = Drug Abuse Screening Tool |  |  |  |  |

Medical clearance for participants to be enrolled will be made by study physicians and staff using the study eligibility criteria, including negative results on the TMS Adult Safety Screen (TASS), MRI Safety and OSU-TBI screening forms. Concurrent medications will be documented as per the latest TMS guidelines [29].

To increase scientific rigor, participants will be asked to provide exhaled breath CO levels at baseline, quit date, end-of-treatment, and at 3-month follow-up visits. Breath CO levels are an effective way of monitoring for cessation, and have an additional positive impact on shaping and inducing smoking abstinence in the first week of a quit attempt [30]. We will use CO monitors for bioverification [including the use of mobile device-based (video) exhaled CO monitoring] to determine smoking abstinence based on a standardized cutoff value of < 5 ppm [31]. This cut-off value has previously been identified as optimal [32], with good sensitivity and specificity among outpatients [33]. In addition, at the 3-month follow-up visits, urinary cotinine will also be collected for more comprehensive bioverification.

### Treatment Selection

There will be two treatment conditions, active-iTBS + CBT + NRT (*n*=25) and sham-iTBS + CBT + NRT (*n*=25). Participants will be randomized by a member of the study team who is not directly involved in administrating study procedures using a simple block design (*n*=4) to minimize imbalances in group allocation. Participants will be randomized into one of two groups, which will be allocated to receive either active- or sham-iTBS. All other team members will remain blinded to treatment conditions throughout study activities. Unblinding will not be permissible unless obligated by the Data Monitoring Committee (DMC).

### Treatments

#### Smoking Cessation Counseling

Five weekly evidence-based CBT for smoking cessation sessions [10] will be provided by a therapist with at least a Master’s degree in mental health (e.g., psychology, social work), and who is rigorously trained and receiving ongoing supervision. The counseling sessions will be performed remotely via telephone except for the Quit Date session, which will be provided in-person. See Appendix 7 for copies of the treatment manuals. At each study visit, including counseling sessions, the Timeline Follow-Back (TLFB) method will be used to gather daily reports of smoking [34].

#### Smoking Cessation Pharmacotherapy

All eligible participants will be prescribed NRT [nicotine patch and at least one rescue method (nicotine gum or lozenge)] to begin on the participant’s intended quit date and continue for at least 60 days. NRT will be prescribed by the study physician, who will work with each participant’s primary care physician or psychiatrist to discuss any contraindications. If no contact with the primary care physician can be made, the participant’s health information will be evaluated by the study physician, who will determine medical clearance to participate in the trial. As mentioned above, neither varenicline nor bupropion will be prescribed due to potential for confounding or influencing seizure risk.

#### MRI Procedures

Participants will undergo an initial MRI scan (pre-iTBS MRI) to obtain structural imaging (T1-weighted scan) and fMRI using a blood oxygen level dependent (BOLD) signal while eyes are open, providing rsFC. The target site for iTBS will be determined for each participant by transforming the location of the right postcentral gyrus cluster that is most positively connected to the posterior insula according to voxel-wise resting-state functional connectivity (see analysis section for details). This personalized cortical stimulation target site will then be uploaded to the neuronavigation system, which will be used in real time to guide the spatial targeting of iTBS. Following the last iTBS session, a second structural and functional MRI scan (post-iTBS MRI) will be conducted at least 1 hour and up to 24 hours later. The stimulation target site will be determined for each participant by transforming the location of the postcentral gyrus cluster functionally connected to the posterior insula into individual anatomical space (using the pre-iTBS anatomical MRI scan), then drawing an 8 × 8 × 8 mm cubed region of interest (ROI) over the cluster location (approximate Montreal Neurological Institute coordinates: 64, −5, 28). Resting state functional images will be preprocessed using freely available open-source statistical parametric mapping software, SPM12 [35]. Functional images will be slice-time corrected and temporally realigned. Individual anatomical images will be co-registered to the functional images and the anatomical images segmented into gray matter, white matter, and cerebral spinal fluid and normalized to the International Consortium for Brain Mapping (ICBM) template. Images will be resliced to 2 × 2 × 2 mm voxels and spatially smoothed using a 5-mm full width at half-maximum Gaussian kernel. Subject-level rsFC will be analyzed using the CONN toolbox [36].

#### Cue Provocation Procedure

Immediately prior to the administration of iTBS, a cue provocation procedure will be implemented in which participants watch a 6-minute video of smoking-related images and follow prompts to engage with a cigarette and lighter. Similar to previous studies using rTMS for the treatment of TUD, activation of neurocircuitry related to addiction by provocation makes it more amenable to modulation [37]. Developed during the pilot project, 60 tobacco-related images including photos of people smoking were extracted from the cross-validated SmoCuDa database of smoking cues [38] based on highest rankings for “urge to smoke” on a 100-point visual analogue scale (overall mean ± SD = 46.75 ± 27.27) as well as images containing persons of color, veterans, and those with common smoking triggers (e.g., alcohol, coffee, multiple people smoking). Instructions to remove a cigarette and lighter out of a plastic bag, place the cigarette on the lips without lighting it, and then put it back into the bag are repeated 4 times. Participants will be asked to complete questionnaires on levels of cravings before and after the provocation procedure as well as after iTBS.

#### TMS Procedures

Administration of rTMS will be performed using a MagVenture MagPro X100 and a Cool-B65 A/P figure-of-eight coil. Individual rMT will be determined prior to sham-controlled rTMS and used to calibrate the intensity of treatment. To determine rMT, electrodes (Neuroline 720, Ambu) will be placed on the first dorsal interosseous (FDI) muscle of the left hand in a belly-tendon montage, and motor evoked potentials (MEPs) will be recorded by an electromyogram (EMG; Power Lab and LabChart) while single-pulse TMS is administered at the motor “hot spot,” defined as the position over the right motor cortex that elicits the greatest MEP in the left first FDI muscle. Resting motor threshold is then determined by the TMS pulse intensity producing on average a MEP of 50 μV peak-to-peak amplitude, using a maximum likelihood estimator (TMS Motor Threshold Assessment Tool, MTAT 2.0) [39]. TMS coil placement for treatment sessions will then be guided in real time by a Brainsight neuronavigational system (Rogue Research) using infrared cameras to co-register participants’ pre-determined stimulation target site with the locations of the head and TMS coil. The transformed ROI in the individual’s MRI space will be utilized to determine the optimal TMS coil setup [40] that maximizes the induced ROI electric field [41]. In more detail, ROI E-field will be maximized perpendicular in direction with respect to the nearest sulci wall. The optimal coil placement will then be loaded into Brainsight as the desired coil setup for the TMS coil operator. iTBS will be administered twice daily separated by 50 minutes for 5 consecutive days. iTBS consists of burst of three TMS pulses at 50 Hz, each pulse at intensity of 90% rMT at cortex, bursting at 5 Hz frequency for a 2-second train, with an 8-second inter-train-interval, for 1800 total pulses (approximately 9 minutes duration) per session. A checklist will be used throughout study administration to promote protocol fidelity between study team members.

#### Outcomes

Feasibility endpoints have been operationalized to allow for quantitative assessment. Feasibility will be measured by evaluating recruitment and retention of participants, with a target enrollment of 50 participant and 80% retention rate (i.e., 40 participants completing at least 90% of study procedures). Acceptability will be measured using a 10-point scale assessing satisfaction of the study procedures (TMS, CBT, MRI, etc.); the TMS intervention will be considered acceptable with an average score of ≥7/10.

We will estimate effect size by evaluating the 7-day point prevalence of smoking based on subject self-report and confirmed using exhaled CO level at end-of-treatment and 3-months post-quit date with abstinence rates averaged over all assessments to take advantage of all available data and adjust for within-subjects clustering of data and using previously established methods of self-reported abstinence and bioverification. Non-abstinence will be defined as self-reported smoking (or other tobacco use) for 7 consecutive days or at least once a week for 2 consecutive weeks. Participants will be considered abstinent if they meet certain criteria. For participants taking NRT at the time of follow-up, they will be considered abstinent if they self-report prolonged abstinence and provide a CO reading < 5 ppm. For those not taking NRT, abstinence will be based on 1) self-reported prolonged abstinence, 2) absence of any biochemical samples indicating smoking (CO ≥ 5 ppm or cotinine ≥ 6 ng/mL), and 3) presence of at least one biochemical sample indicating abstinence (CO < 5 ppm and/or cotinine < 6 ng/mL).

Comparisons of continuous demographic and clinical data will be completed with linear regressions. Comparisons of dichotomous demographic and clinical data will use generalized linear models using a binomial distribution. Regarding 7-day point prevalence smoking abstinence outcomes at end-of-treatment, and 3-months, odds ratios and 95% confidence intervals will be computed from estimates obtained in generalized linear models. A longitudinal analysis using generalized linear models will also be conducted on abstinence across end-of-treatment and 3-month time periods with predictors of treatment arm, time in months (standardized and centered), and the treatment arm-by-time interaction. All missing smoking data will be replaced as non-abstinent (i.e., missing = smoking), so that participants with missing follow-up data will be assumed to be smoking. All analyses will be conducted in R version 4.0.4. No interim analyses are planned.

Secondary outcomes will include daily number of cigarettes smoked, subjective levels of withdrawal, craving, and urges to smoke, 7-day point prevalence at 3-months post-quit date, as well as smoking relapse, which is defined as smoking 5 or more cigarettes per day for 3 consecutive days. We will evaluate 7-day point prevalence smoking abstinence by calculating a dichotomous outcome variable (abstinent or not abstinent) to determine the proportion of participants with bioverified abstinence at each assessment from quit date through 3 months follow-up. Due to the relatively small sample size, we will describe the rates of smoking abstinence by calculating the proportion of participants who are confirmed to be abstinent. We will calculate confidence intervals around the effect size to determine the level of variability expected in the subsequent full RCT.

For the neuroimaging data, movement and scan artifacts will be detected using Artifact Detection Tools (National Imaging Tools and Resources Collaboratory) and included in the rsFC analyses as covariates of no interest. Group-level analyses will be conducted in SPM12. rsFC analyses will be confined to a bilateral insula regional of interest (ROI) mask defined from the Automated Anatomical Labeling (AAL) atlas with a significance threshold of p < .001 uncorrected, cluster extent > 10 voxels. Exploratory whole brain analyses will also be conducted with a significance threshold of p < .001 uncorrected, cluster extent > 50 voxels. For neural target engagement, we will evaluate functional connectivity between the rTMS target at the post-central gyrus and the right posterior insula. We will also evaluate the correlation between changes on rsFC pre- and post-rTMS and change in the number of cigarettes smoked in the past 24 hours. We will also use a data-driven approach by employing group independent component analysis (ICA) to compare functional network connectivity (FNC) between active- and sham-rTMS groups including FC between rTMS target and right posterior insula.

Regarding safety, we will evaluate for adverse events (AEs) at each study visit. rTMS-specific adverse events will be evaluated at the beginning and end of each TMS session using the rTMS Acute Side Effects Questionnaire (**Table 2**). Side effects to NRT will be assessed at each visit following the initiation of NRT at the Quit Date session. Suicide risk assessment is conducted at each counseling session. The PI will meet at least weekly with study personnel to discuss enrollment, participation, and any adverse events or unanticipated problems. Regular meetings between investigators and the project manager will allow for ongoing progress reports, including the number of participants currently involved in each study, attrition rates, and scheduled data collection from participants, as well as notification and review of any AEs. Safety monitoring for AEs will be conducted in real time by the PI and/or project manager. The following information about adverse events will be collected: 1) the onset and resolution of the AE, 2) an assessment of the severity or intensity (use existing grading scales whenever possible), 3) an assessment of the relationship of the event to the study (definitely, probably, possibly or not related), and 4) action taken (e.g., none, referral to physician, start or increase concomitant medication). The PI will determine the severity of the event, will assign attribution to the event, and will monitor the event until its resolution. Any adverse events will be reported to the DVAHCS and DUMC IRBs in accordance with their Human Research Protection Program’s Standards of Practice. All research projects conducted at DVAHCS and DUMC are required to have yearly IRB review, including a safety review. Additionally, any changes to the project between review periods must be approved by the appropriate IRB prior to fielding. A VA Clinical Science Research and Development (CSRD) DMC has been assigned to oversee the safety of the study. The DMC is primarily responsible for safeguarding the interests of study participants, assessing the safety and efficacy of trial interventions, and monitoring the progress of the study. The DMC will conduct reviews of the clinical trial and monitor recruitment every 12 months. This DMC serves as an independent advisory group to the CSRD Director and provides recommendations about proceeding with the study initially, continuing the study at subsequent reviews, or stopping the study. The DMC is responsible for identifying mechanisms for the completion of various tasks that will impact the safety and efficacy of study procedures and overall conduct of the study. All Serious Adverse Events (SAEs) and Unanticipated Problem Involving Risks to Subjects or Others (UPIRTSOs) will be submitted to the DMC within 3 business days. AEs will be reported in aggregate within progress reports.

This study, formally titled “Neuroimaging correlates and feasibility of transcranial magnetic stimulation (TMS) to improve smoking cessation outcomes in veterans with comorbid PTSD” (TMS-STOP), has received approvals from the Institutional Review Boards (IRBs) of DUMC (Pro00107816) and DVAHCS (1625460). The trial plans to begin enrollment in July 2023 at a single study site (DVAHCS). Information about enrollment status can be found at ClinicalTrials.gov (NCT05723588). Data will be managed and protected in accordance with the research data and safety standards of practice at DVAHCS and DUMC. To ensure confidentiality, all records will be identified by the participant’s identification number, not by name. All raw hard copy data will be kept in a locked file cabinet in a locked room on DVAHCS property. Data files will be stored in a limited access folder on a secure server to which only study staff members have access.

## Discussion

This RCT will investigate the feasibility, acceptability, and efficacy of fMRI-guided, accelerated iTBS combined with CBT and NRT protocol among a target subject population of veterans with PTSD. This multimodal intervention is particularly novel as it is the first focal rTMS protocol for smoking cessation targeting the insula through functional connectivity while integrating psychotherapy and NRT. The feasibility of this treatment approach has been investigated in an open-label trial. Preliminary outcomes suggest that this approach is acceptable, feasible, and may suggest efficacy for the veterans with PTSD who have participated thus far [17]. Strengths of this study as compared to our recently completed pilot study are the addition of a sham-control group, modification of rTMS to a more efficient patterned form of stimulation (iTBS), incorporation of an accelerated treatment schedule (twice daily treatments), as well as the inclusion of a 3-month post-quit date follow up visit to evaluate longer-term outcomes.

This protocol offers several innovations to previous studies and could have major implications on the future treatment of veterans with PTSD. The outcomes of this study will provide preliminary evidence that a focal TMS smoking cessation intervention has evidence for feasibility, acceptability, and efficacy for veterans with PTSD. If successful, further clinical research will be required to replicate the results in an adequately powered phase III RCT. Ultimately, this clinical intervention could be incorporated into smoking cessation and interventional specialty VA clinics as a part of the National VA Clinical rTMS Program. Such a treatment option could enhance available treatments for smoking cessation at VA and elsewhere. Additional studies of this treatment protocol may be adapted to veterans without PTSD, as well as for civilian populations.

While this study protocol has important strengths, there are weaknesses. Strict eligibility criteria will allow enrollment of the target patient population of veterans with PTSD. However, as noted in our pilot study, a consequence of this is that only about one-half of the participants we screened were able to fit the stringent eligibility criteria. This study is limited to individuals who smoke cigarettes, and so future studies will need to also evaluate the treatment’s applicability to other forms of nicotine consumption (e.g., vaping). Furthermore, while individualized TMS targeting using functional connectivity to the insula is novel, the feasibility and cost-effectiveness of these procedures in a clinical setting will need to be further evaluated. It is possible that the technical steps required to utilize this protocol including rsFC determination, optimal coil placement, and navigated TMS would exceed the capacity of most clinical rTMS programs. It will be important to consider modifications that simplify and automate the technical steps to make it more feasible to translate this intervention into the clinical setting.

Finally, there are many future directions that can be explored following the results of this study. For instance, assuming positive outcomes, we intend to organize a multi-center phase III RCT with sufficient power to conclusively determine efficacy of the intervention. Additionally, further explorations within the parameter space may enhance treatment efficacy. These could include increasing the number of sessions, adjusting TMS parameters for improved target engagement, or re-evaluating the timing of the various interventions relative to planned quit dates.

## Conclusion

Many veterans struggle with PTSD and TUD. This randomized control trial, guided by a recently completed open-label trial, will help determine feasibility, acceptability, and effectiveness of treating smoking cessation with a combination of TMS, CBT and NRT. Overall, this study fills a critical gap as foundational research to better inform larger studies which may help improve treatment options for individuals with TUD, and eventually may help inform decisions about service coverages and reimbursement.

## Data Availability

No datasets were generated or analysed during the current study. All relevant data from this study will be made available upon study completion.

## References

1. Cornelius ME, L.C., Jamal A, et al., Tobacco Product Use Among Adults – United States, 2021. MMWR Morb Mortal Wkly Rep 2023. 72:475**–**483.

2. Mokdad, A.H., et al., Actual causes of death in the United States, 2000. JAMA, 2004. 291(10): p. 1238–45.

3. Centers for Disease, C. and Prevention, Annual smoking-attributable mortality, years of potential life lost, and economic costs--United States, 1995-1999. MMWR Morb Mortal Wkly Rep, 2002. 51(14): p. 300–3.

4. Sokal, J., et al., Comorbidity of medical illnesses among adults with serious mental illness who are receiving community psychiatric services. J Nerv Ment Dis, 2004. 192(6): p. 421–7.

5. Joseph, A.M., et al., Smoking intensity and severity of specific symptom clusters in posttraumatic stress disorder. J Trauma Stress, 2012. 25(1): p. 10–6.

6. Tam, J., K.E. Warner, and R. Meza, Smoking and the Reduced Life Expectancy of Individuals With Serious Mental Illness. Am J Prev Med, 2016. 51(6): p. 958–966.

7. Beckham, J.C., et al., Predictors of lapse in first week of smoking abstinence in PTSD and non-PTSD smokers. Nicotine Tob Res, 2013. 15(6): p. 1122–9.

8. Dedert, E.A., et al., A Randomized Clinical Trial of Nicotine Preloading for Smoking Cessation in People with Posttraumatic Stress Disorder. J Dual Diagn, 2018. 14(3): p. 148–157.

9. Fiore, M.C., Treating tobacco use and dependence: an introduction to the US Public Health Service Clinical Practice Guideline. Respir Care, 2000. 45(10): p. 1196–9.

10. McFall, M., et al., Integrating tobacco cessation into mental health care for posttraumatic stress disorder: a randomized controlled trial. JAMA, 2010. 304(22): p. 2485–93.

11. Addicott, M.A., et al., Increased Functional Connectivity in an Insula-Based Network is Associated with Improved Smoking Cessation Outcomes. Neuropsychopharmacology, 2015. 40(11): p. 2648–56.

12. BrainsWay. BrainsWay Receives FDA Clearance for Smoking Addiction in Adults. 2020; Available from: https://www.brainsway.com/news_events/brainsway-receives-fda-clearance-for-smoking-addiction-in-adults/.

13. Hauer, L., et al., Effects of repetitive transcranial magnetic stimulation on nicotine consumption and craving: A systematic review. Psychiatry Res, 2019. 281: p. 112562.

14. Blumberger, D.M., et al., Effectiveness of theta burst versus high-frequency repetitive transcranial magnetic stimulation in patients with depression (THREE-D): a randomised non-inferiority trial. Lancet, 2018. 391(10131): p. 1683–1692.

15. Cole, E.J., et al., Stanford Neuromodulation Therapy (SNT): A Double-Blind Randomized Controlled Trial. Am J Psychiatry, 2022. 179(2): p. 132–141.

16. Addicott, M.A., et al., Low- and High-Frequency Repetitive Transcranial Magnetic Stimulation Effects on Resting-State Functional Connectivity Between the Postcentral Gyrus and the Insula. Brain Connect, 2019. 9(4): p. 322–328.

17. Young JR, G.J., Polick C, Papanikolas W, Evans M, Dedert E, Addicott M, Moore SD, Appelbaum LG, Beckham JC., Multimodal Smoking Cessation Using rTMS, CBT and NRT in Veterans with PTSD: Results from a Nonrandomized Phase II Trial, in Society of Biological Psychiatry (SOBP). 2023: San Diego, CA.

18. Gorgolewski, K.J. and R.A. Poldrack, A Practical Guide for Improving Transparency and Reproducibility in Neuroimaging Research. PLoS Biol, 2016. 14(7): p. e1002506.

19. Heatherton, T.F., et al., The Fagerstrom Test for Nicotine Dependence: a revision of the Fagerstrom Tolerance Questionnaire. Br J Addict, 1991. 86(9): p. 1119–27.

20. Cox, L.S., S.T. Tiffany, and A.G. Christen, Evaluation of the brief questionnaire of smoking urges (QSU-brief) in laboratory and clinical settings. Nicotine Tob Res, 2001. 3(1): p. 7–16.

21. Hughes, J.R. and D. Hatsukami, Signs and symptoms of tobacco withdrawal. Arch Gen Psychiatry, 1986. 43(3): p. 289–94.

22. Weathers, F.W., Litz, B.T., Keane, T.M., Palmieri, P.A., Marx, B.P., & Schnurr, P.P., The PTSD Checklist for DSM-5 (PCL-5). Scale available from the National Center for PTSD *at* www.ptsd.va.gov. 2013.

23. Beck, A.T., Steer, R. A., & Brown, G., Beck Depression Inventory–II (BDI-II). APA PsycTests, 1996.

24. Bush, K., et al., The AUDIT alcohol consumption questions (AUDIT-C): an effective brief screening test for problem drinking. Ambulatory Care Quality Improvement Project (ACQUIP). Alcohol Use Disorders Identification Test. Arch Intern Med, 1998. 158(16): p. 1789–95.

25. Staley, D. and N. el-Guebaly, Psychometric properties of the Drug Abuse Screening Test in a psychiatric patient population. Addict Behav, 1990. 15(3): p. 257–64.

26. Association, F.a.D. FDA Drug Safety Communication: FDA updates label for stop smoking drug Chantix (varenicline) to include potential alcohol interaction, rare risk of seizures, and studies of side effects on mood, behavior, or thinking. 2018; Available from: https://www.fda.gov/drugs/drug-safety-and-availability/fda-drug-safety-communication-fda-updates-label-stop-smoking-drug-chantix-varenicline-include.

27. Little, M.A. and J.O. Ebbert, The safety of treatments for tobacco use disorder. Expert Opin Drug Saf, 2016. 15(3): p. 333–41.

28. Keel, J.C., M.J. Smith, and E.M. Wassermann, A safety screening questionnaire for transcranial magnetic stimulation. Clin Neurophysiol, 2001. 112(4): p. 720.

29. Rossi, S., et al., Safety and recommendations for TMS use in healthy subjects and patient populations, with updates on training, ethical and regulatory issues: Expert Guidelines. Clin Neurophysiol, 2021. 132(1): p. 269–306.

30. Benowitz, N.L., J. Hukkanen, and P. Jacob, 3rd, *Nicotine chemistry, metabolism, kinetics and biomarkers*. Handb Exp Pharmacol, 2009(192): p. 29–60.

31. Beckham, J.C., et al., Mobile contingency management as an adjunctive treatment for co-morbid cannabis use disorder and cigarette smoking. Addict Behav, 2018. 79: p. 86–92.

32. Woodward, M. and H. Tunstall-Pedoe, An iterative technique for identifying smoking deceivers with application to the Scottish Heart Health Study. Prev Med, 1992. 21(1): p. 88–97.

33. Middleton, E.T. and A.H. Morice, Breath carbon monoxide as an indication of smoking habit. Chest, 2000. 117(3): p. 758–63.

34. Lewis-Esquerre, J.M., et al., Validation of the timeline follow-back in the assessment of adolescent smoking. Drug Alcohol Depend, 2005. 79(1): p. 33–43.

35. Neuroiminaging, W.C.f.H. Statistical parametric mapping software, SP*M12*. 2020; Available from: https://www.fil.ion.ucl.ac.uk/spm/software/spm12/.

36. Whitfield-Gabrieli, S. and A. Nieto-Castanon, Conn: a functional connectivity toolbox for correlated and anticorrelated brain networks. Brain Connect, 2012. 2(3): p. 125–41.

37. Zangen, A., et al., Repetitive transcranial magnetic stimulation for smoking cessation: a pivotal multicenter double-blind randomized controlled trial. World Psychiatry, 2021. 20(3): p. 397–404.

38. Manoliu, A., et al., SmoCuDa: A Validated Smoking Cue Database to Reliably Induce Craving in Tobacco Use Disorder. Eur Addict Res, 2021. 27(2): p. 107–114.

39. F., A. and B. J. TMS Motor Threshold Assessment Tool 2.0 (MTAT 2.0). 2011; Available from: https://www.clinicalresearcher.org/software.htm.

40. Dannhauer, M., et al., TAP: targeting and analysis pipeline for optimization and verification of coil placement in transcranial magnetic stimulation. J Neural Eng, 2022. 19(2).

41. Gomez, L.J., M. Dannhauer, and A.V. Peterchev, Fast computational optimization of TMS coil placement for individualized electric field targeting. Neuroimage, 2021. 228: p. 117696.

